# Development and validation of composite inflammaging metrics: Findings from two prospective cohort studies

**DOI:** 10.1101/2023.05.13.23289903

**Authors:** Chenxi Li, Yumeng Ge, Zhenqing Yang, Gan Yang, Xingqi Cao, Jinjing Fu, Zuyun Liu

## Abstract

**Background:** Aging-related inflammation is associated with chronic diseases and mortality. This study aimed to: 1) develop composite inflammaging metrics (CIMs) in UK biobank (UKB), and validate them in UKB and National Health and Nutrition Examination Survey (NHANES); 2) estimate mortality and CVD risk predictions of CIMs; 3) compare CIMs with single inflammatory blood biomarkers and conventional inflammatory indexes; 4) examine associations between lifestyles and CIMs.

**Methods:** We utilized algorithms including multiple linear regression, principal component analysis (PCA), allostatic load (AL), and Klemera and Doubal method (KDM), to develop four CIMs from five inflammatory blood biomarkers, using data of 438,321 adults (40-70 years) from UK Biobank (UKB). We validated these CIMs in UKB and 10,667 adults (20-84 years) from NHANES IV. We performed a parametric proportional hazard model based on Gompertz distribution to estimate CVD and mortality risk predictions of CIMs. Areas under receiver operating characteristic curves (AUCs) were calculated to compare the predictive abilities of CIMs. Multiple linear regression models were used to access associations between lifestyles and CIMs.

**Results:** With adjustment for age and sex, four CIMs were significantly associated with higher risks of all-cause mortality and incident CVD in UKB, among which CIM_KDM_ outperformed the others (all-cause mortality: hazard ratio [HR] = 1.48, 95% confidence interval [CI] = 1.46, 1.50; incident CVD: HR = 1.34, 95% CI = 1.33, 1.36). CIM_KDM_ had the best discriminative ability for predicting 10-year survival and incident CVD in UKB (all-cause mortality: AUC = 0.728; incident CVD: AUC = 0.712). CIMs were responsive to lifestyle variables. For example, in UKB, compared to never smokers, current smokers had a significant increment in CIM_KDM_ (coefficient = 0.30 SD, *P* < 0.001). Similar results were well validated in NHANES IV.

**Conclusions:** We developed and validated four novel CIMs that were predictive of mortality and CVD risk. CIM_KDM_ outperformed the others and had the potential to be used in aging related preventive and intervention programs. Intervention programs targeting lifestyles could slow inflammaging and further reduce disease burden.

## 1. Introduction

Inflammation has been recognized as one of the important factors driving aging^1^. Low-grade, chronic, and systemic inflammation has been termed as “inflammaging” in aging process, which can result in high risk of mortality and age-related chronic disease^2-4^, such as cardiovascular disease (CVD)^5^, and Alzheimer’s disease^6^. Previous evidence has revealed that commonly used acute inflammation biomarkers, e.g., interleukin-6 (IL-6), C-reactive protein (CRP), and tumor necrosis factor-alpha (TNF-α), can independently capture the effect of chronic inflammation on mortality^7,8^. There are still no canonical standard biomarkers for inflammaging^11^. Moreover, one certain inflammatory biomarker cannot comprehensively mirror the whole inflammatory status in the aging process, which emphasizes the importance of composite measures^12^.

Considering the necessity to identify the underlying inflammaging states, researchers have developed lots of composite inflammatory indexes as predictors of mortality and chronic diseases. Of conventional composite inflammatory indexes, systemic inflammation response index (SIRI)^14^, systemic immune-inflammation index (SII)^15^, neutrophil-to-lymphocyte ratio (NLR) ^16^, platelet-to-lymphocyte ratio (PLR) ^17^, and lymphocyte-to-monocyte ratio (LMR)^18^ have been widely applied in predicting mortality and pre-diagnostic age-related disease^20-22^. Besides, some researchers have used advanced algorithms, such as principal component analysis (PCA)^23^ and deep-learning^24^, trying to construct inflammaging metrics, but these metrics are too complex, and lack generalization in diverse population of wide age range. Therefore, there is a need to develop new inflammaging metric with practical value.

While Klemera and Doubal method (KDM) has been widely used in developing biological aging measure^25-27^, it has rarely been utilized in inflammatory estimation yet. Besides, allostatic load (AL) is usually regarded as an indicator of physiological dysregulation resulting from cumulative physiological stress that in relation to health outcomes along aging^28-30^, of which inflammation strain is a portion. In this study, four algorithms including multiple linear regression, PCA, AL, and KDM were used to develop composite inflammaging metrics (CIMs). Afterwards, it is vital to verify whether these CIMs have a better performance than single inflammatory blood biomarkers and conventional inflammatory indexes. Slowing inflammaging process through modifiable factors is a natural need for public health. Whether healthy lifestyles could influence inflammaging status reflected by CIMs is significant. Thus, this study aimed to: 1) develop four CIMs in UK biobank (UKB), and validate them in UKB and National Health and Nutrition Examination Survey (NHANES); 2) estimate mortality and CVD risk predictions of CIMs; 3) compare CIMs with single inflammatory blood biomarkers and conventional inflammatory indexes; 4) examine associations between lifestyles and CIMs.

## 2. Methods

### 2.1 Study participants

UKB is a large prospective cohort of national participants from across the UK, which recruited over 500,000 participants from 2006 to 2010 at 22 assessment centers.

Participants’ information on demographics, lifestyle, medical conditions, and other health-related aspects was provided through interviews and physical measurements at baseline. More detailed information on UKB can be seen elsewhere^31^. We excluded participants with missing data on covariates (N = 23,243) or five inflammatory blood biomarkers (N = 40,882), leaving 438,321 participants aged 40-70 years to develop and validate CIMs. Details of the participants selection in UKB are shown in **Figure S1**. UKB study was approved by the North West Multi-centre Research Ethics Committee (ref. 11/NW/0382). All participants provided written informed consent.

NHANES is an ongoing programme aiming to evaluate US civilian’s health and nutritional status, carried out by the National Center for Health Statistics. NHANES collected data from nationally representative cross-sectional surveys. More detailed information on NHANES can be seen elsewhere (https://www.cdc.gov/nchs/nhanes/index.htm). We used data from NHANES IV (1999–2010, N = 13,823) as an independent validation sample. We excluded participants who did not accomplish 8 hours of fasting before blood sampling or those with missing data on biomarkers (N = 1,368), follow-up time (N = 15), survey weights (N = 1,008), or covariates (N = 765). We included 10,667 participants aged 20-84 years in final analytic sample to validate CIMs. Details of participants selection in NHANES IV are shown in **Figure S1**. NHANES was approved by the National Center for Health Statistics Research Ethics Review Board. All participants provided informed consent.

### 2.2 Inflammatory metrics

#### 2.2.1 Inflammatory blood biomarkers

Considering the feasibility and accessibility of novel inflammaging metric, we included five blood biomarkers: platelet count (1000cell/uL), lymphocyte count (1000cell/uL), monocyte count (1000cell/uL), neutrophil count (1000cell/uL), and CRP (g/L). Lymphocyte count, monocyte count, neutrophil count, and CRP were logarithmically transformed to approximate normal distribution in regression.

#### 2.2.2 Conventional inflammatory indexes

We calculated five conventional inflammatory indexes based on inflammatory blood biomarkers: SII^15^, SIRI^14^, NLR^32,33^, PLR^17,34^, and LMR^18,35^. The formula is presented as follows, where P, N, L, and M are platelet, neutrophil, lymphocyte, and monocyte counts, respectively:

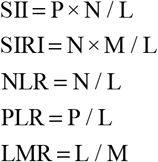

#### 2.2.3 CIM_lm_ (composite inflammaging metric using linear regression)

We used multiple linear regression model to regress five inflammatory blood biomarkers on age, and defined prediction as CIM_lm_. The formula of CIM_lm_ is presented as follows:

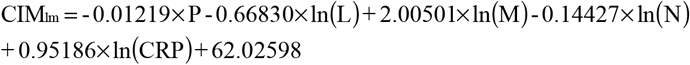

#### 2.2.4 CIM_PCA_ (composite inflammaging metric using Principal Component Analysis)

We did PCA with five inflammatory blood biomarkers, and took four components out of five according to Scree Plot (**Figure S2**) and cumulative proportion of variance interpretation (over 80%, in **Table S1**). Then, we used multiple linear regression model to regress four selected components on age, and defined prediction as CIM_PCA_. The formula of CIM_PCA_ is presented as follows:

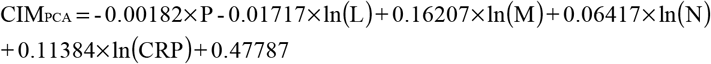

#### 2.2.5 CIM_AL_ (composite inflammaging metric using allostatic load)

We used AL^29^ to calculate CIM_AL_ based on five inflammatory blood biomarkers. The 75th percentile (25th percentile additionally for platelet count) was determined as the cut-off for five biomarkers. Every biomarker with values above the cut-off (below additionally for platelet count) was defined as ‘1’ otherwise ‘0’, and the sum of five biomarkers was calculated as CIM_AL_, ranging from 0 to 5. The higher CIM_AL_, the worse a participant’s inflammatory status. The normal range (defined as ‘0’) for five inflammatory blood biomarkers is presented as follows:

1. 213.4-287.0 1000cell/uL for P;
2. 0-2.30 1000cell/uL for L;
3. 0-0.57 1000cell/uL for M;
4. 0-4.95 1000cell/uL for N;
5. 0-2.72 g/L for CRP.

#### 2.2.6 CIM_KDM_ (composite inflammaging metric using Klemera-Doubal Method)

We used KDM^25,36^ to calculate CIM_KDM_ based on five inflammatory blood biomarkers. The formula of CIM_KDM_ is presented as follows:

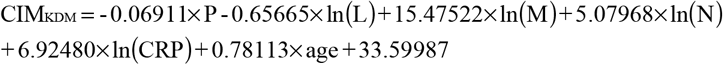

### 2.3 Outcomes

The primary outcome in UKB was all-cause mortality, which was obtained from the National Health Service Central Register Scotland (Scotland) and the National Health Service Information Centre (England and Wales). Detailed information is available from https://content.digital.nhs.uk/services. In this study, participants were followed up from baseline to the date of death or Aug 29, 2021 (censoring time), whichever occurred first. The secondary outcome in UKB was incident CVD events. We defined CVD according to the International Statistical Classification of Diseases and Related Health Problems 9^th^ (ICD-9) codes (410.x, 411.x, 412.x, 413.x, 414.x,429.79, 430.x, 431.x, 432.x, 433.x, 434.x, 435.x, 436.x, 437.x, 438.x), and 10^th^ edition (ICD-10) codes (I20.x, I21.x, I22.x, I23.x, I24.1, I25.x, I46, I60.x, I61.x, I63.x, I64.x)^37^.

Participants who were diagnosed as CVD at baseline were excluded. Participants were followed up from baseline to the first diagnosis of CVD, the date of death, or Aug 29, 2021(censoring time), whichever occurred first.

The primary outcome in NAHNES IV was all-cause mortality. NHANES IV data were linked to the National Death Index, provided by the Centers for Disease Control and Prevention (https://www.cdc.gov/nchs/data-linkage/mortality-public.htm). Participants were followed up from baseline to the date of death or December 31, 2019 (censoring time), whichever occurred first. The secondary outcome in NAHNES IV was CVD-specific mortality. We defined CVD-specific mortality using ICD-10 codes (I00-I09, I11, I13, I20-I51, or I60-I69)^38^.

### 2.4 Covariates and lifestyles

In UKB, covariates included age, sex (male and female), ethnic group (white and others), educational level (high, intermediate, and low), current employment status (working, retired, and others), Townsend deprivation index^39^, and disease counts (ranging 0-9, including dementia, CVD, hypertension, cancer, diabetes, cholelithiasis, osteoarthritis, respiratory disease, and depression). In NHANES IV, covariates included age, sex (male and female), ethnic group (non-Hispanic white, non-Hispanic black, Hispanic, and others), educational level (high, intermediate, and low), and disease counts (ranging 0-10, including congestive heart failure, stroke, cancer, chronic bronchitis, emphysema, cataracts, arthritis, type 2 diabetes mellitus, hypertension, and myocardial infraction).

In both UKB and NHANES IV, lifestyles included smoking status (never, previous, and current), alcohol intake frequency (“never or special occasions only”, “one to three times per month”, “one to four times per week”, and “daily or almost daily”), regular exercise (yes and no)^39,40^, healthy diet (yes and no)^39,40^, and body mass index (BMI) (<18.5, ≥18.5 and <25, ≥25 and <30, and ≥30 kg/m^2^).

### 2.5 Statistical analyses

Characteristics of the participants in two analytic samples were presented as numbers (percentages) for categorical variables, and mean ± standard deviation (SD) for continuous variables, respectively. All inflammatory metrics were scaled in analyses to assure them comparable.

First, we developed four CIMs in UKB. Second, we examined whether CIMs could stratify the risk of outcomes onset. We observed incident event rate plotted against CIMs percentiles. We plotted Kaplan-Meier curves of cumulative hazard of outcomes for persons in the lowest 20%, the middle 20% and the highest 20% of CIMs. Third, we estimated CIMs’ predictive validity in mortality and incident CVD with a basic model including age and sex. We used a parametric proportional hazard model (Gompertz distribution) to evaluate the associations between CIMs and outcomes, and estimated hazard ratios (HRs). We estimated areas under receiver operating characteristic curves (AUCs) of 10-year outcomes incidence risk prediction. We used calibration curve to observe CIMs’ discriminative performance on 10-year outcomes incidence risk predictions. Fourth, we estimated HRs and AUCs for single inflammatory blood biomarkers and conventional inflammatory indexes to examine whether CIMs had a better performance than them. Sixth, we used multiple linear regression model to evaluate associations between lifestyles and inflammatory metrics with all covariates adjusted for, to observe whether healthy lifestyles can slow a person’s inflammaging status. Seventh, we calculated four CIMs in NHANES IV and repeated all the analyses with outcomes as all-cause mortality and CVD-specific mortality. Eighth, we did two sensitivity analyses of estimation on HRs and AUCs: 1) in relatively healthy population (disease counts equal to 0); 2) with disease counts additionally adjusted for.

We used R version 4.1 to conduct all statistical analyses. A *P* value < 0.05 (two-tailed) was defined as statistically significant.

## 3. Results

### 3.1 Characteristics of study participants

Characteristics of participants were presented in **Table S2**. Of 438,321 participants in UKB, the mean age was 57.0 ± 8.1 years, and 200,788 (45.8%) participants were male. A total of 29,218 (6.7%) participants died during a median follow-up time of 12.1 years (interquartile range, IQR 11.4–12.8 years), and a total of 31,812 (7.8%) participants out of 409,460 (excluding 28,861 participants diagnosed with CVD at baseline) developed CVD during a median follow-up time of 12.4 years (IQR 11.6– 13.2 years).

Of 10,667 participants in the NHANES IV, the mean age was 48.9 ± 17.7 years, and 5,265 (49.4%) participants were male. A total of 2,047 (19.2%) participants died during a median follow-up time of 13.1 years (IQR 10.3–17.0 years), and a total of 608 (5.7%) participants developed CVD-specific death during the same follow-up time.

### 3.2 Characteristics of CIMs

In UKB, mean of CIM_lm_ was 57.0 years (SD = 1.4). Mean of CIM_PCA_ was 0.0 (SD = 0.2). Mean of CIM_AL_ was 1.5 (SD = 1.2). Mean of CIM_KDM_ was 56.5 years (SD = 13.3). In NHANES IV, mean of CIM_lm_ was 57.6 years (SD = 1.5). Mean of CIM_PCA_ was 0.0 (SD = 0.2). Mean of CIM_AL_ was 1.8 (SD = 1.2). Mean of CIM_KDM_ was 54.7 years (SD = 19.5). **Figure S3** presents CIMs’ distribution. **Figure S4** presents CIMs’ correlation with age. As expected, CIM_KDM_ were highly correlated with age, because age was in CIM_KDM_ measure. Other CIMs showed weak correlation with age.

### 3.3 CIMs stratify the risk of outcomes onset

As shown in **Figure 1a**, we observed increasing all-cause mortality and CVD incidence rates over CIM percentiles in UKB. As shown in **Figure 1b**, we observed increasing all-cause mortality and CVD-specific mortality rates roughly over CIM percentiles in NHANES IV. CIM_KDM_ outperformed the other.

**Figure 1.**
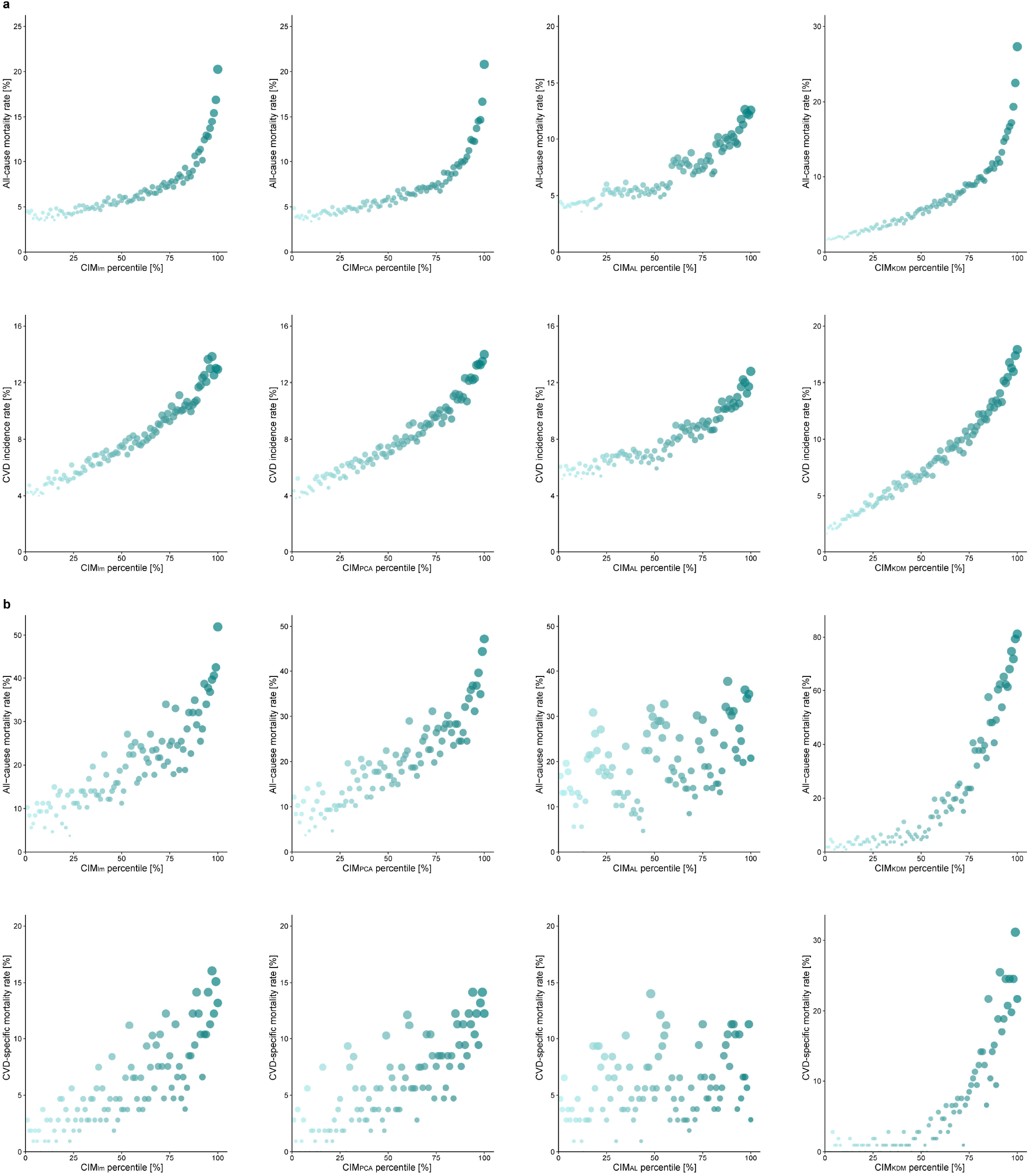
Associations between CIMs and event rate in UK Biobank and NHANES IV. **a**. All-cause mortality rate and CVD incidence rate based on CIM percentile in UK Biobank. **b**. All-cause mortality rate and CVD-specific mortality rate based on CIM percentile in NHANES IV. Abbreviation: NHANES, National Health and Nutrition Examination Survey; CIM, Composite inflammaging metric; CIM_lm_; Composite inflammaging metric using linear regression; CIM_PCA_, Composite inflammaging metric using Principal Component Analysis (PCA); CIM_AL_ Composite inflammaging metric using allostatic load; CIM_KDM_, Composite inflammaging metric using Klemera-Doubal Method (KDM).

We calculated the cumulative hazard for persons in the lowest 20%, the middle 20% and the highest 20% of CIMs (**Figure 2**), with 95% confidence intervals (CIs). As shown in **Figure 2a**, all CIMs stratified the different risk level of outcomes onset clearly in UKB, among which CIM_KDM_ outperformed the other. As shown in **Figure 2b**, CIM_lm_ and CIM_PCA_ stratified the different risk level of outcomes onset clearly in NHANES IV. CIM_AL_ barely stratify the risk level of persons in the middle 20% and the highest 20%, while CIM_KDM_ barely stratify the risk level of persons in the middle 20% and the lowest 20%.

**Figure 2.**
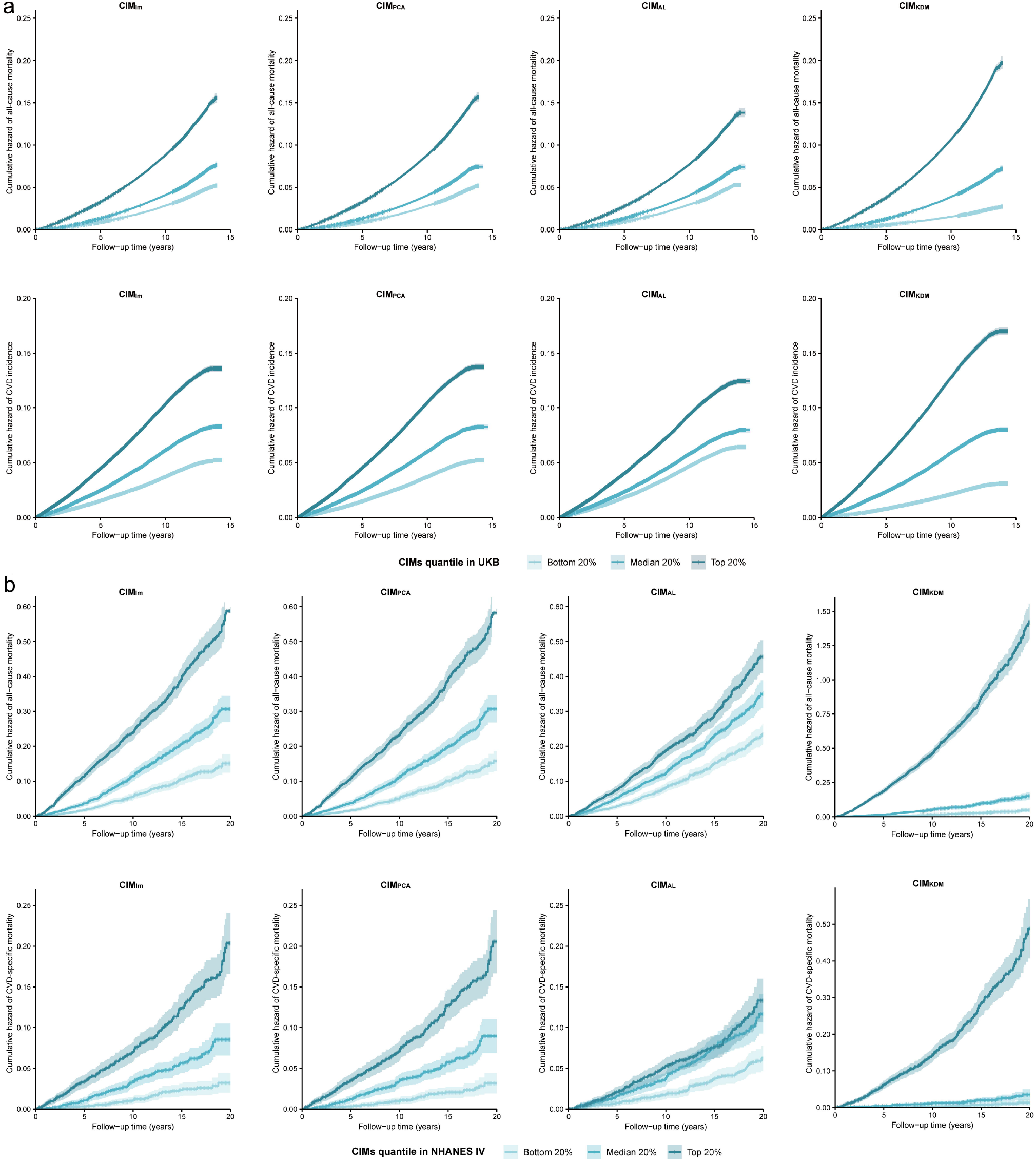
Cumulative hazard curves of event rate in UK Biobank and NHANES IV. **a**. Cumulative hazard of all-cause mortality and CVD incidence in UK Biobank stratified by CIMs quantiles. **b**. Cumulative hazard of all-cause mortality and CVD-specific mortality in NHNAES IV stratified by CIMs quantiles. CIMs quantiles are indicated by light green (bottom 20%), green (median 20%), dark green (top 20%) with 95% confidence intervals. Abbreviations: NHANES, National Health and Nutrition Examination Survey; CIM, Composite inflammaging metric; CIM_lm_; Composite inflammaging metric using linear regression; CIM_PCA_, Composite inflammaging metric using Principal Component Analysis (PCA); CIM_AL_ Composite inflammaging metric using allostatic load; CIM_KDM_, Composite inflammaging metric using Klemera-Doubal Method (KDM).

### 3.4 Associations between inflammatory metrics and outcomes

**Table 1** shows the associations between inflammatory metrics and outcomes with age and sex adjusted for. We found that in UKB, each 1-SD increase in CIM_lm_ increased the risk of all-cause mortality and CVD incidence by 32% (HR = 1.32, 95% CI = 1.30, 1.33) and 22% (HR = 1.22, 95% CI = 1.20, 1.23). Each 1-SD increase in CIM_PCA_ increased the risk of all-cause mortality and CVD incidence by 33% (HR = 1.33, 95% CI = 1.31, 1.34) and 23% (HR = 1.23, 95% CI = 1.22, 1.25). Each 1-SD increase in CIM_AL_ increased the risk of all-cause mortality and CVD incidence by 33% (HR = 1.33, 95% CI = 1.31, 1.34) and 23% (HR = 1.23, 95% CI = 1.22, 1.24). Each 1-SD increase in CIM_KDM_ increased the risk of all-cause mortality and CVD incidence by 48% (HR = 1.48, 95% CI = 1.46, 1.50) and 34% (HR = 1.34, 95% CI = 1.33, 1.36).

**Table 1.**
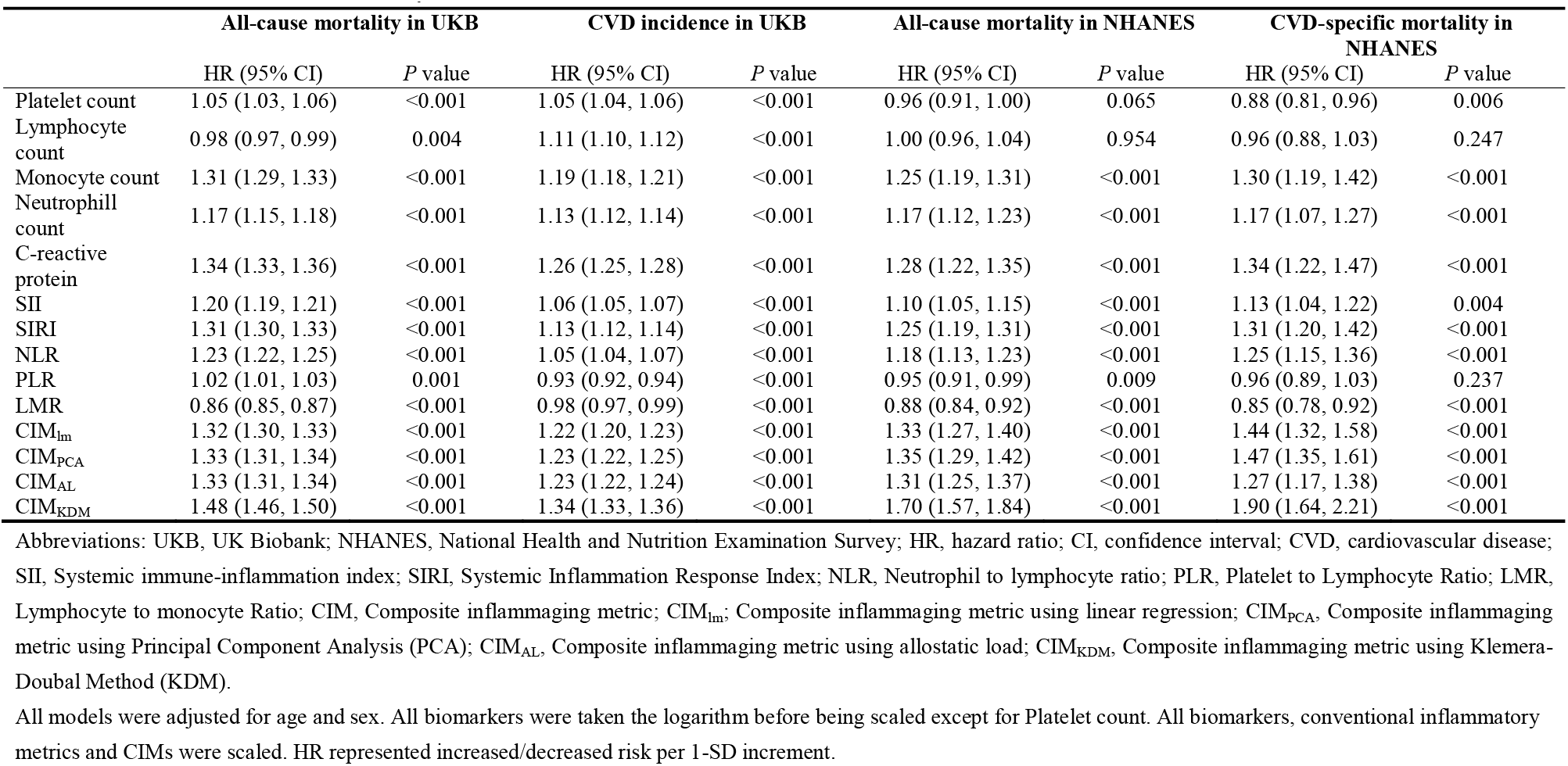
Associations between inflammatory metrics and outcomes.

|  | All-cause mortality in UKB |  | CVD incidence in UKB |  | All-cause mortality in NHANES |  | CVD-specific mortality in NHANES |  |
| --- | --- | --- | --- | --- | --- | --- | --- | --- |
|  | HR (95% CI) | *P* value | HR (95% CI) | *P* value | HR (95% CI) | *P* value | HR (95% CI) | *P* value |
| Platelet count | 1.05 (1.03, 1.06) | <0.001 | 1.05 (1.04, 1.06) | <0.001 | 0.96 (0.91, 1.00) | 0.065 | 0.88 (0.81, 0.96) | 0.006 |
| Lymphocyte count | 0.98 (0.97, 0.99) | 0.004 | 1.11 (1.10, 1.12) | <0.001 | 1.00 (0.96, 1.04) | 0.954 | 0.96 (0.88, 1.03) | 0.247 |
| Monocyte count | 1.31 (1.29, 1.33) | <0.001 | 1.19 (1.18, 1.21) | <0.001 | 1.25 (1.19, 1.31) | <0.001 | 1.30 (1.19, 1.42) | <0.001 |
| Neutrophil count | 1.17 (1.15, 1.18) | <0.001 | 1.13 (1.12, 1.14) | <0.001 | 1.17 (1.12, 1.23) | <0.001 | 1.17 (1.07, 1.27) | <0.001 |
| C-reactive protein | 1.34 (1.33, 1.36) | <0.001 | 1.26 (1.25, 1.28) | <0.001 | 1.28 (1.22, 1.35) | <0.001 | 1.34 (1.22, 1.47) | <0.001 |
| SII | 1.20 (1.19, 1.21) | <0.001 | 1.06 (1.05, 1.07) | <0.001 | 1.10 (1.05, 1.15) | <0.001 | 1.13 (1.04, 1.22) | 0.004 |
| SIRI | 1.31 (1.30, 1.33) | <0.001 | 1.13 (1.12, 1.14) | <0.001 | 1.25 (1.19, 1.31) | <0.001 | 1.31 (1.20, 1.42) | <0.001 |
| NLR | 1.23 (1.22, 1.25) | <0.001 | 1.05 (1.04, 1.07) | <0.001 | 1.18 (1.13, 1.23) | <0.001 | 1.25 (1.15, 1.36) | <0.001 |
| PLR | 1.02 (1.01, 1.03) | 0.001 | 0.93 (0.92, 0.94) | <0.001 | 0.95 (0.91, 0.99) | 0.009 | 0.96 (0.89, 1.03) | 0.237 |
| LMR | 0.86 (0.85, 0.87) | <0.001 | 0.98 (0.97, 0.99) | <0.001 | 0.88 (0.84, 0.92) | <0.001 | 0.85 (0.78, 0.92) | <0.001 |
| CIM<sub>lm</sub> | 1.32 (1.30, 1.33) | <0.001 | 1.22 (1.20, 1.23) | <0.001 | 1.33 (1.27, 1.40) | <0.001 | 1.44 (1.32, 1.58) | <0.001 |
| CIM<sub>PCA</sub> | 1.33 (1.31, 1.34) | <0.001 | 1.23 (1.22, 1.25) | <0.001 | 1.35 (1.29, 1.42) | <0.001 | 1.47 (1.35, 1.61) | <0.001 |
| CIM<sub>AL</sub> | 1.33 (1.31, 1.34) | <0.001 | 1.23 (1.22, 1.24) | <0.001 | 1.31 (1.25, 1.37) | <0.001 | 1.27 (1.17, 1.38) | <0.001 |
| CIM<sub>KDM</sub> | 1.48 (1.46, 1.50) | <0.001 | 1.34 (1.33, 1.36) | <0.001 | 1.70 (1.57, 1.84) | <0.001 | 1.90 (1.64, 2.21) | <0.001 |

Abbreviations: UKB, UK Biobank; NHANES, National Health and Nutrition Examination Survey; HR, hazard ratio; CI, confidence interval; CVD, cardiovascular disease; SII, Systemic immune-inflammation index; SIRI, Systemic Inflammation Response Index; NLR, Neutrophil to lymphocyte ratio; PLR, Platelet to Lymphocyte Ratio; LMR, Lymphocyte to monocyte Ratio; CIM, Composite inflammaging metric; CIM<sub>lm</sub>, Composite inflammaging metric using linear regression; CIM<sub>PCA</sub>, Composite inflammaging metric using Principal Component Analysis (PCA); CIM<sub>AL</sub>, Composite inflammaging metric using allostatic load; CIM<sub>KDM</sub>, Composite inflammaging metric using Klemere-Doubal Method (KDM).
All models were adjusted for age and sex. All biomarkers were taken the logarithm before being scaled except for Platelet count. All biomarkers, conventional inflammatory metrics and CIMs were scaled. HR represented increased/decreased risk per 1-SD increment.

Among single inflammatory blood biomarkers, CRP performed the best. Each 1-SD increase in ln(CRP) increased the risk of all-cause mortality and CVD incidence by 34% (HR = 1.34, 95% CI = 1.33, 1.36) and 26% (HR = 1.26, 95% CI = 1.25, 1.28).

Among conventional inflammatory indexes, SIRI performed the best. Each 1-SD increase in SIRI increased the risk of all-cause mortality and CVD incidence by 31% (HR = 1.31, 95% CI = 1.30, 1.33) and 13% (HR = 1.13, 95% CI = 1.12, 1.14).

Overall, CIMs had a better performance than single inflammatory blood biomarkers and conventional inflammatory indexes, among which CIM_KDM_ outperformed the other. Similar results were found in NHANES IV as outcomes were all-cause mortality and CVD-specific mortality.

Figure 3. shows AUCs with a basic model including age and sex. All models of CIMs well calibrated in UKB and NHANES IV for 10-year event rate (**Figure S5**). For 10-year all-cause mortality and CVD incidence in UKB, basic model had an AUC of 0.713 (95% CI = 0.709, 0.716) and 0.700 (95% CI = 0.696, 0.703). CIM_lm_ had an AUC of 0.725 (95% CI = 0.721, 0.728) and 0.708 (95% CI = 0.705, 0.711). CIM_PCA_ had an AUC of 0.725 (95% CI = 0.722, 0.729) and 0.709 (95% CI = 0.706, 0.712).

CIM_AL_ had an AUC of 0.728 (95% CI = 0.724, 0.731) and 0.710 (95% CI = 0.707, 0.713). CIM_KDM_ had an AUC of 0.728 (95% CI = 0.725, 0.732) and 0.712 (95% CI = 0.709, 0.715). Among single inflammatory blood biomarkers, CRP performed the best. ln(CRP) had an AUC of 0.728 (95% CI = 0.725, 0.731) and 0.712 (95% CI = 0.709, 0.715). Among conventional inflammatory indexes, SIRI performed the best. SIRI had an AUC of 0.724 (95% CI = 0.721, 0.728) and 0.703 (95% CI = 0.700, 0.706).

**Figure 3.**
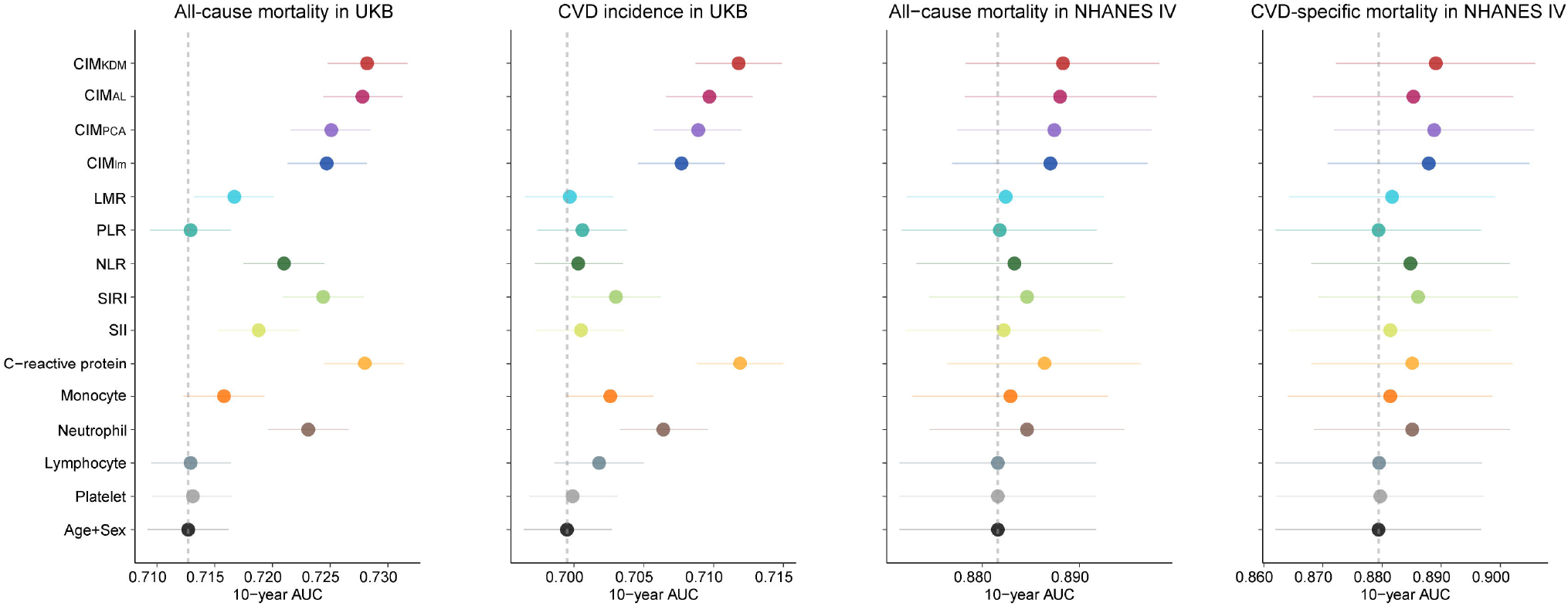
The discrimination performance of biomarkers, conventional inflammatory metrics and CIMs in UK Biobank and NHANES IV. This figure shows the predictive ability of inflammatory metrics for 10-year all-cause mortality, 10-year CVD incidence in UKB and NHANES IV. The dotted line indicates a basic model, Age+Sex. All models were adjusted for age and sex. Data is shown as mean AUC (center dot of error bar) and 95% confidence interval (line of error bar). Abbreviations: NHANES, National Health and Nutrition Examination Survey; AUC, area under the curve; SII, Systemic immune-inflammation index; SIRI, Systemic Inflammation Response Index; NLR, Neutrophil to lymphocyte ratio; PLR, Platelet to Lymphocyte Ratio; LMR, Lymphocyte to monocyte Ratio; CIM, Composite inflammaging metric; CIM_lm_; Composite inflammaging metric using linear regression; CIM_PCA_, Composite inflammaging metric using Principal Component Analysis (PCA); CIM_AL_ Composite inflammaging metric using allostatic load; CIM_KDM_, Composite inflammaging metric using Klemera-Doubal Method (KDM).

CIMs had a better performance than conventional inflammatory indexes. CIM_KDM_ and CRP had similar great performance in UKB. However, in NHANES IV, CIM_KDM_ outperformed CRP. For 10-year all-cause mortality and CVD-specific mortality, ln(CRP) had an AUC of 0.886 (95% CI = 0.876, 0.896) and 0.885 (95% CI = 0.868, 0.902). CIM_KDM_ had an AUC of 0.888 (95% CI = 0.878, 0.898) and 0.889 (95% CI = 0.872, 0.906). Compared to single inflammatory blood biomarkers and conventional inflammatory indexes, CIMs had a better performance in NHANES IV.

### 3.5 Associations between lifestyles with inflammatory metrics

Figure 4. shows the associations between all lifestyle variables (including smoking status, alcohol intake frequency, regular exercise, healthy diet and BMI, simultaneously input) with inflammatory metrics.

Overall, inflammatory blood biomarkers, conventional inflammatory indexes, and CIMs were responsive to lifestyle variables. Unhealthy lifestyles were associated with a serious inflammatory status. For example, in UKB, compared to never smokers, current smokers had a significant increment in ln(CRP) (coefficient = 0.28 SD, *P* < 0.001), SII (coefficient = 0.05 SD, *P* < 0.001), and CIM_KDM_ (coefficient = 0.30 SD, *P* < 0.001). Interestingly, two relatively unhealthy lifestyle statuses (high alcohol intake frequency and BMI < 18.5) were associated with lower level of inflammatory blood biomarkers and CIMs. For example, in UKB, compared to participants with alcohol intake frequency of “never or special occasions only”, participants with alcohol intake frequency of “daily or almost daily” had a significant decrement in ln(CRP) (coefficient = -0.10 SD, *P* < 0.001) and CIM_KDM_ (coefficient = -0.04 SD, *P* < 0.001). Compared to participants with normal weight (BMI ≥ 18.5 and < 25), underweight participants (BMI < 18.5) had a significant decrement in ln(CRP) (coefficient = -0.50 SD, *P* < 0.001) and CIM_KDM_ (coefficient = -0.27 SD, *P* < 0.001). The results in NHANES IV were similar to those in UKB.

**Figure 4.**
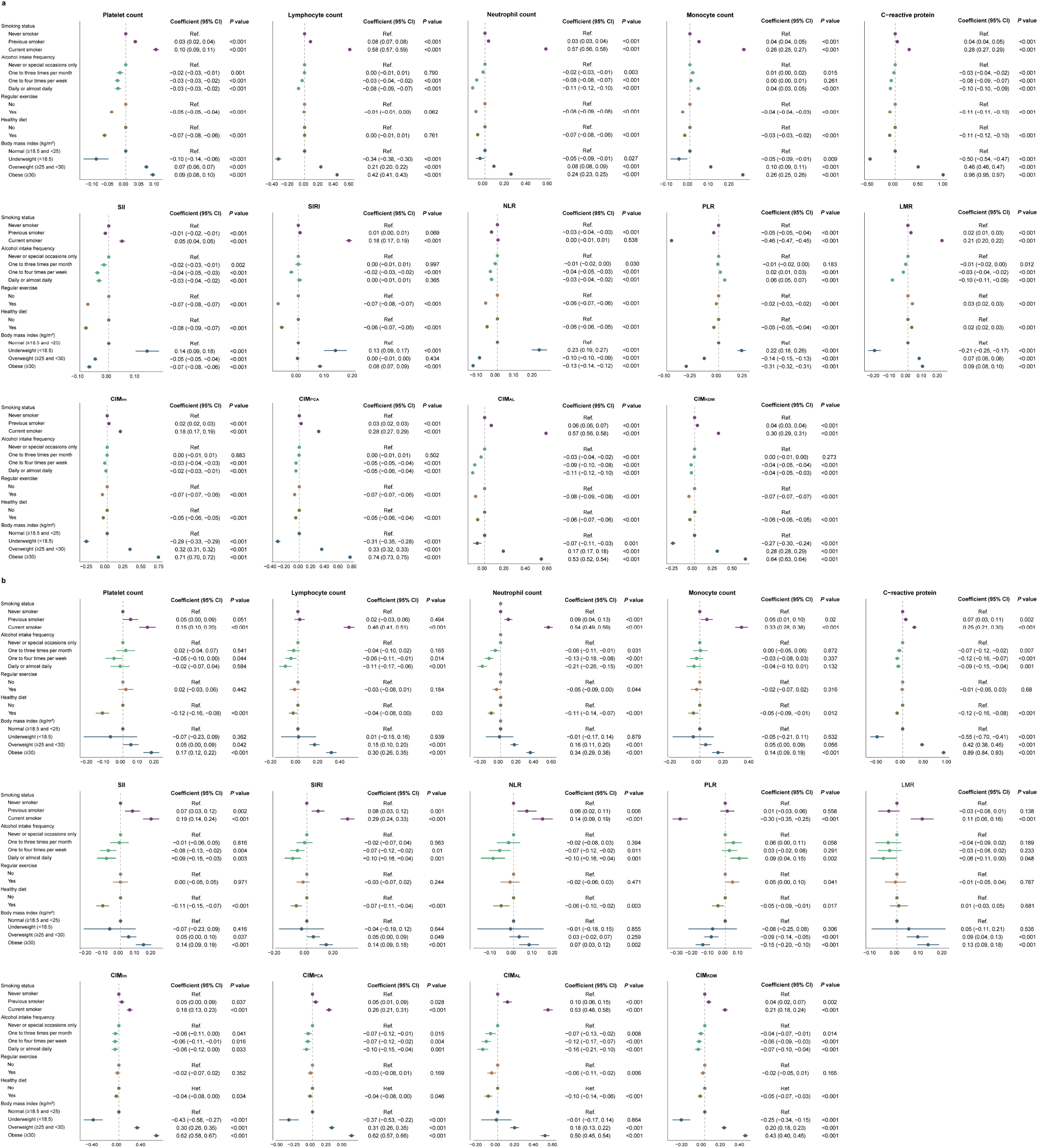
Associations between inflammatory metrics and lifestyles in UKB and NHANES IV. **a**. Models were adjusted for age, sex, ethnic group, education, current employment status, Townsend deprivation index, and disease counts to examine the associations of indicators with lifestyles in UKB. **b**. Models were adjusted for age, sex, ethnic group, education, and disease counts to examine the associations of indicators with lifestyles in NHANES IV. Abbreviations: NHANES, National Health and Nutrition Examination Survey; SII, Systemic immune-inflammation index; SIRI, Systemic Inflammation Response Index; NLR, Neutrophil to lymphocyte ratio; PLR, Platelet to Lymphocyte Ratio; LMR, Lymphocyte to monocyte Ratio; CIM, Composite inflammaging metric; CIM_lm_; Composite inflammaging metric using linear regression; CIM_PCA_, Composite inflammaging metric using Principal Component Analysis (PCA); CIM_AL_ Composite inflammaging metric using allostatic load; CIM_KDM_, Composite inflammaging metric using Klemera-Doubal Method (KDM).

### 3.6 Sensitivity analyses

We repeated analyses of estimation on HRs and AUCs twice under conditions below: 1) in relatively healthy population (disease counts equal to 0) (**Table S3**); 2) with disease counts additionally adjusted for (**Table S4**). We observed no substantial changes in results.

## 4. Discussion

In this large-scale population study, we developed and validated novel measures of inflammaging status—CIMs, including CIM_lm_, CIM_PCA_, CIM_AL_ and CIM_KDM_. These metrics were demonstrated to be leveraged as predictors of all-cause mortality, CVD incidence, and CVD-specific mortality. It is observed that CIMs capturing risk more precisely beyond single inflammatory blood biomarkers and conventional inflammatory indexes, with exception of CRP. Furthermore, CIM_KDM_ outperformed other CIMs in that it successfully stratified risk and manifested its preponderance of discrimination and calibration in 10-year outcomes predictions. The association between some common unhealthy lifestyles and lower CIMs elucidated the feasibility of deferring inflammaging process by lifestyle intervention.

Our metrics took five inflammatory biomarkers into consideration, that is platelet count, lymphocyte count, monocyte count, neutrophil count, and CRP, which are commonly tested in clinical practice. As we noticed, platelet count, lymphocyte count or monocyte count were not stable enough to be an indicator, which might be attributed to the complex mechanism of immune system. In the case of lymphocyte, T cells are deemed to be the main modulatory participants in atherosclerotic which accounts for the majority of CVD; yet the diverse differentiations and possible transformation^12^ of T lymphocytes determine their function to secrete pro- or anti-inflammatory cytokines, thus explaining why both the promotive and protective effects coexist^41-43^. With that, some studies^44,45^ held the viewpoint that low lymphocyte count poses threat to CVD occurrence and progression. Though we assumed composite metrics would surpass individual biomarkers, the good performance of CRP could not be denied as its performance was close to CIM_KDM_.

Of conventional inflammatory indexes, elevated SIRI, having the similar capability to assess the risk of mortality with CIM_lm_, seemed to be stronger predictors than others. Compared to these former indexes, in principle, CIMs had more edge over the prediction of onset across CVD than of mortality risk. On the one hand, conventional inflammatory indexes were calculated by multiplication and division among the platelet count, lymphocyte count, monocyte count, and neutrophil count, whereas the supposed functions of lymphocyte and monocyte were contradictory. According to our discussion above, it is untenable to posit that one certain inflammatory biomarker stay consistencies in its physiological roles of inflammaging. On the other hand, none of these indexes considered CRP while it is certainly an important inflammatory biomarker. Therefore, CIMs (CIM_KDM_ in particular) are better than conventional inflammatory indexes from the perspective of algorithm. Moreover, conventional inflammatory indexes are regularly used, and might more suitable for linking systemic inflammation to prognosis or adverse outcomes in patients with cancer, CVD^46,47^, and so on. For instance, our findings suggested the probability of the negative correlations between PLR/LMR and outcomes. Similarly, another prospective study^48^ based on UKB also reported inconsistent results that there are positive associations with risk of several cancers for PLR but negative associations for LMR.

To the best of our knowledge, we are the first to derive inflammaging metrics from platelet count, lymphocyte count, monocyte count, neutrophil count, and CRP, plus via four algorithms in such a nationwide prospective cohort. There are discrepancies between CIMs in predicting mortality and CVD risk. CIM_KDM_ was the most pronounced one since it was robust in participants who were relatively healthy at baseline. Additionally, the strength of CIM_KDM_ was conspicuous in drawing a distinction between high and low risk groups and higher accuracy of prediction. While the discriminatory power of CIM_lm_ and CIM_PCA_ were inferior to CIM_KDM_, they could also be valuable metrics for CVD risk prediction since they predicted with sufficient accuracy as well as CIM_KDM_. The hazard trajectory of all-cause mortality and CVD-specific mortality in NHANES IV suggested that, when follow-up time became longer, CIM_KDM_ was plausible of less predictive value for individual of moderate risk. Still, sample size was also an inevitable factor to this deficiency. Even though, as an excellent biological aging measures proved by many former investigations^26,36^, KDM remains its intrinsic superiority of algorithm over multiple linear regression and PCA accordingly. Such a overlap of hazard trajectory emerged in different quantile of CIM_AL_ likewise. CIM_AL_ could be a fine indicator in terms of stability and discrimination, but its ability to identify people at risk was limited inasmuch as AL score is a discrete variable. More concretely, CIM_AL_ was an integer ranged between 0 to 5, which meant the possible score of it was much smaller versus widespread AL score that combined more biomarkers from various systems^28,49^.

Inflammaging has been perceived as a vital target for antiaging and curbing the onset or progression of age-related disease^50^. Chronic exposure to exogenous risk factors like unhealthy lifestyles^51^, augments inflammatory burden risk, thereby fueling inflammaging^5^. Many researches on pathophysiology have already revealed that unhealthy lifestyles, such as smoking^52,53^, alcohol intake^54^, physical inactivity^55,56^, unhealthy diet^57,58^, and obesity^59,60^, could trigger or increase chronic inflammation. At same time, associations of unhealthy lifestyles with CVD and mortality risk was widely illustrated in cohort studies^61,62^. Therefore, with evaluation of the association of inflammaging indexes and unhealthy lifestyles, we found that those who kept away from smoking, did regular exercise, had healthy diet and kept fit, had lower CIMs. That gave us an insight into healthy lifestyles that may well attenuate inflammaging. However, alcohol intake and underweight were shown as positive factors to CIMs. Alcohol intake was assumed to have a J-shaped relationship between CVD, namely low-to-moderate drinkers have lower risk^63^ since some beverage types like red wine are likely to exert anti-inflammatory and antioxidant effects^64^. Thus, while moderate amount of alcohol intake remains controversial, our study offered some evidence for the relationship between inflammatory biomarkers and alcohol intake. Obesity is accompanied by chronic, low-grade inflammation^60^; but the concomitant low fat-free mass in the aging process has been linked to inflammation as well, for example, the cachexia in CVD^65^ or cancer^66^. According to a previous study, middle-aged people’s BMI varied linearly with CRP while elderly people with low body mass showed chronic inflammation^67^. So, a possible explanation was that most of the underweight participants were not being undernourished. Broadly speaking, positively alteration of unhealthy lifestyles would contribute to practical approaches to prevention and intervention of inflammaging and related diseases.

The major strength of our study is that we derived CIMs via four well-performed algorithms in a large-scale prospective cohort (i.e., UKB) and achieved external validation in another independent population-based cohort (i.e., NHANES). The generalization of results in sensitivity analyses were basically recognized. Besides, five primary inflammatory biomarkers used by CIMs are of quick availability, easy detectability and low-priced examination cost. Meanwhile, there are several limitations in our study. Firstly, given the small sample size of the validation sample (i.e., NHANES), general transferability of CIMs needs to be validated in larger cohorts. Secondly, to enhance the prediction ability, the coverage of biomarkers calls for extension, and more inflammaging biomarkers with practical application value need to be identified. Finally, in sight of complicated mechanisms on pathophysiology, advanced methods that incorporating two opposite aspects of inflammation and immune may be favorable for representing inflammaging^23^. Of course, more aetiological researches are expected to offer deeper interpretation to inflammaging.

## Data Availability

NHANES data produced are available online at (https://www.cdc.gov/nchs/nhanes/index.htm).
UKB data has been conducted using the UKB resource under application number 61856.

## Conclusions

In summary, applying four algorithms—multiple linear regression, PCA, AL, and KDM, we developed four novel CIMs in UKB and validated them externally in US adults. Furthermore, we elucidated their value in mortality and incident CVD risk prediction, and verified their response to modifiable lifestyle factors. The findings support the implementation of preventive strategies and intervention programs targeting lifestyles to slow inflammaging and further reduce disease burden.

## Acknowledgments

This study has been conducted using the UK Biobank resource under application number 61856. We wish to thank the UK Biobank participants providing the sample that made the data available. We also thank all participants who took part in the National Health and Nutrition Examination Survey.

## Author contributions

Z.L. designed and supervised the study. C.L. and Y.G. analyzed the data. C.L., Y.G., and Z.L. interpreted the results. C.L. and Y.G. drafted the manuscript. All authors revised the manuscript. Z.L. took responsibility for the content of the article. All authors read and approved the final version of the manuscript.

## Reference

1. Kennedy BK, Berger SL, Brunet A, et al. Geroscience: linking aging to chronic disease. Cell. 2014;159(4):709–713.

2. Franceschi C, Bonafe M, Valensin S, et al. Inflamm-aging. An evolutionary perspective on immunosenescence. Ann N Y Acad Sci. 2000;908:244–254.

3. Franceschi C, Campisi J. Chronic inflammation (inflammaging) and its potential contribution to age-associated diseases. J Gerontol A Biol Sci Med Sci. 2014;69 Suppl 1:S4–9.

4. Franceschi C, Garagnani P, Parini P, Giuliani C, Santoro A. Inflammaging: a new immune-metabolic viewpoint for age-related diseases. Nat Rev Endocrinol. 2018;14(10):576–590.

5. Liberale L, Montecucco F, Tardif JC, Libby P, Camici GG. Inflamm-ageing: the role of inflammation in age-dependent cardiovascular disease. Eur Heart J. 2020;41(31):2974–2982.

6. Newcombe EA, Camats-Perna J, Silva ML, Valmas N, Huat TJ, Medeiros R. Inflammation: the link between comorbidities, genetics, and Alzheimer’s disease. J Neuroinflammation. 2018;15(1):276.

7. Varadhan R, Yao W, Matteini A, et al. Simple biologically informed inflammatory index of two serum cytokines predicts 10 year all-cause mortality in older adults. J Gerontol A Biol Sci Med Sci. 2014;69(2):165–173.

8. Giovannini S, Onder G, Liperoti R, et al. Interleukin-6, C-reactive protein, and tumor necrosis factor-alpha as predictors of mortality in frail, community-living elderly individuals. J Am Geriatr Soc. 2011;59(9):1679–1685.

9. Alberro A, Iribarren-Lopez A, Saenz-Cuesta M, Matheu A, Vergara I, Otaegui D. Inflammaging markers characteristic of advanced age show similar levels with frailty and dependency. Sci Rep. 2021;11(1):4358.

10. Michaud M, Balardy L, Moulis G, et al. Proinflammatory cytokines, aging, and age-related diseases. J Am Med Dir Assoc. 2013;14(12):877–882.

11. Furman D, Campisi J, Verdin E, et al. Chronic inflammation in the etiology of disease across the life span. Nat Med. 2019;25(12):1822–1832.

12. Fulop T, Cohen A, Wong G, Witkowski JM, Larbi A. Are There Reliable Biomarkers for Immunosenescence and Inflammaging? In: Moskalev A, ed. Biomarkers of Human Aging. Cham: Springer International Publishing; 2019:231–251.

13. Ligthart S, Vaez A, Vosa U, et al. Genome Analyses of >200,000 Individuals Identify 58 Loci for Chronic Inflammation and Highlight Pathways that Link Inflammation and Complex Disorders. Am J Hum Genet. 2018;103(5):691–706.

14. Qi Q, Zhuang L, Shen Y, et al. A novel systemic inflammation response index (SIRI) for predicting the survival of patients with pancreatic cancer after chemotherapy. Cancer. 2016;122(14):2158–2167.

15. Hu B, Yang XR, Xu Y, et al. Systemic immune-inflammation index predicts prognosis of patients after curative resection for hepatocellular carcinoma. Clinical cancer research : an official journal of the American Association for Cancer Research. 2014;20(23):6212–6222.

16. Cho H, Hur HW, Kim SW, et al. Pre-treatment neutrophil to lymphocyte ratio is elevated in epithelial ovarian cancer and predicts survival after treatment. Cancer Immunol Immunother. 2009;58(1):15–23.

17. Krenn-Pilko S, Langsenlehner U, Thurner EM, et al. The elevated preoperative platelet-to-lymphocyte ratio predicts poor prognosis in breast cancer patients. British journal of cancer. 2014;110(10):2524–2530.

18. Hutterer GC, Stoeckigt C, Stojakovic T, et al. Low preoperative lymphocyte-monocyte ratio (LMR) represents a potentially poor prognostic factor in nonmetastatic clear cell renal cell carcinoma. Urologic oncology. 2014;32(7):1041–1048.

19. Sylman JL, Mitrugno A, Atallah M, et al. The Predictive Value of Inflammation-Related Peripheral Blood Measurements in Cancer Staging and Prognosis. Front Oncol. 2018;8:78.

20. Xu M, Chen R, Liu L, et al. Systemic immune-inflammation index and incident cardiovascular diseases among middle-aged and elderly Chinese adults: The Dongfeng-Tongji cohort study. Atherosclerosis. 2021;323:20–29.

21. Li H, Wu X, Bai Y, et al. Physical activity attenuates the associations of systemic immune-inflammation index with total and cause-specific mortality among middle-aged and older populations. Sci Rep. 2021;11(1):12532.

22. Jin Z, Wu Q, Chen S, et al. The Associations of Two Novel Inflammation Indexes, SII and SIRI with the Risks for Cardiovascular Diseases and All-Cause Mortality: A Ten-Year Follow-Up Study in 85,154 Individuals. J Inflamm Res. 2021;14:131–140.

23. Morrisette-Thomas V, Cohen AA, Fulop T, et al. Inflamm-aging does not simply reflect increases in pro-inflammatory markers. Mech Ageing Dev. 2014;139:49–57.

24. Sayed N, Huang Y, Nguyen K, et al. An inflammatory aging clock (iAge) based on deep learning tracks multimorbidity, immunosenescence, frailty and cardiovascular aging. Nat Aging. 2021;1:598–615.

25. Klemera P, Doubal S. A new approach to the concept and computation of biological age. Mechanisms of ageing and development. 2006;127(3):240–248.

26. Liu Z. Development and Validation of 2 Composite Aging Measures Using Routine Clinical Biomarkers in the Chinese Population: Analyses From 2 Prospective Cohort Studies. J Gerontol A Biol Sci Med Sci. 2021;76(9):1627–1632.

27. Jee H, Park J. Selection of an optimal set of biomarkers and comparative analyses of biological age estimation models in Korean females. Arch Gerontol Geriatr. 2017;70:84–91.

28. Duong MT, Bingham BA, Aldana PC, Chung ST, Sumner AE. Variation in the Calculation of Allostatic Load Score: 21 Examples from NHANES. J Racial Ethn Health Disparities. 2017;4(3):455–461.

29. Seeman TE, Singer BH, Rowe JW, Horwitz RI, McEwen BS. Price of adaptation--allostatic load and its health consequences. MacArthur studies of successful aging. Arch Intern Med. 1997;157(19):2259–2268.

30. Crimmins EM, Johnston M, Hayward M, Seeman T. Age differences in allostatic load: an index of physiological dysregulation. Exp Gerontol. 2003;38(7):731–734.

31. Sudlow C, Gallacher J, Allen N, et al. UK biobank: an open access resource for identifying the causes of a wide range of complex diseases of middle and old age. PLoS Med. 2015;12(3):e1001779.

32. Pichler M, Hutterer GC, Stoeckigt C, et al. Validation of the pre-treatment neutrophil-lymphocyte ratio as a prognostic factor in a large European cohort of renal cell carcinoma patients. British journal of cancer. 2013;108(4):901–907.

33. Hermanns T, Bhindi B, Wei Y, et al. Pre-treatment neutrophil-to-lymphocyte ratio as predictor of adverse outcomes in patients undergoing radical cystectomy for urothelial carcinoma of the bladder. British journal of cancer. 2014;111(3):444–451.

34. Fox P, Hudson M, Brown C, et al. Markers of systemic inflammation predict survival in patients with advanced renal cell cancer. British journal of cancer. 2013;109(1):147–153.

35. Szkandera J, Gerger A, Liegl-Atzwanger B, et al. The lymphocyte/monocyte ratio predicts poor clinical outcome and improves the predictive accuracy in patients with soft tissue sarcomas. International journal of cancer. 2014;135(2):362–370.

36. Levine ME. Modeling the rate of senescence: can estimated biological age predict mortality more accurately than chronological age? The journals of gerontology Series A, Biological sciences and medical sciences. 2013;68(6):667–674.

37. Wan EYF, Fung WT, Schooling CM, et al. Blood Pressure and Risk of Cardiovascular Disease in UK Biobank: A Mendelian Randomization Study. Hypertension. 2021;77(2):367–375.

38. Wan Z, Guo J, Pan A, Chen C, Liu L, Liu G. Association of Serum 25-Hydroxyvitamin D Concentrations With All-Cause and Cause-Specific Mortality Among Individuals With Diabetes. Diabetes care. 2021;44(2):350–357.

39. Chudasama YV, Khunti K, Gillies CL, et al. Healthy lifestyle and life expectancy in people with multimorbidity in the UK Biobank: A longitudinal cohort study. PLoS Med. 2020;17(9):e1003332.

40. Yang Z, Pu F, Cao X, et al. Does healthy lifestyle attenuate the detrimental effects of urinary polycyclic aromatic hydrocarbons on phenotypic aging? An analysis from NHANES 2001-2010. Ecotoxicology and environmental safety. 2022;237:113542.

41. Schwartz DM, Burma AM, Kitakule MM, Luo Y, Mehta NN. T Cells in Autoimmunity-Associated Cardiovascular Diseases. Front Immunol. 2020;11:588776.

42. Hansson GK, Hermansson A. The immune system in atherosclerosis. Nat Immunol. 2011;12(3):204–212.

43. Legein B, Temmerman L, Biessen EA, Lutgens E. Inflammation and immune system interactions in atherosclerosis. Cell Mol Life Sci. 2013;70(20):3847–3869.

44. Shah AD, Denaxas S, Nicholas O, Hingorani AD, Hemingway H. Low eosinophil and low lymphocyte counts and the incidence of 12 cardiovascular diseases: a CALIBER cohort study. Open Heart. 2016;3(2):e000477.

45. Nunez J, Minana G, Bodi V, et al. Low lymphocyte count and cardiovascular diseases. Curr Med Chem. 2011;18(21):3226–3233.

46. Ji Z, Liu G, Guo J, et al. The Neutrophil-to-Lymphocyte Ratio Is an Important Indicator Predicting In-Hospital Death in AMI Patients. Front Cardiovasc Med. 2021;8:706852.

47. Zhai G, Wang J, Liu Y, Zhou Y. Platelet-lymphocyte ratio as a new predictor of in-hospital mortality in cardiac intensive care unit patients. Sci Rep. 2021;11(1):23578.

48. Nost TH, Alcala K, Urbarova I, et al. Systemic inflammation markers and cancer incidence in the UK Biobank. Eur J Epidemiol. 2021;36(8):841–848.

49. Hastings WJ, Almeida DM, Shalev I. Conceptual and Analytical Overlap Between Allostatic Load and Systemic Biological Aging Measures: Analyses From the National Survey of Midlife Development in the United States. J Gerontol A Biol Sci Med Sci. 2022;77(6):1179–1188.

50. Franceschi C, Garagnani P, Vitale G, Capri M, Salvioli S. Inflammaging and ‘Garb-aging’. Trends Endocrinol Metab. 2017;28(3):199–212.

51. Mandelli L, Milaneschi Y, Hiles S, Serretti A, Penninx BW. Unhealthy lifestyle impacts on biological systems involved in stress response: hypothalamic-pituitary-adrenal axis, inflammation and autonomous nervous system. Int Clin Psychopharmacol. 2022.

52. Ambrose JA, Barua RS. The pathophysiology of cigarette smoking and cardiovascular disease: an update. J Am Coll Cardiol. 2004;43(10):1731–1737.

53. Bhalla DK, Hirata F, Rishi AK, Gairola CG. Cigarette smoke, inflammation, and lung injury: a mechanistic perspective. J Toxicol Environ Health B Crit Rev. 2009;12(1):45–64.

54. Bishehsari F, Magno E, Swanson G, et al. Alcohol and Gut-Derived Inflammation. Alcohol Res. 2017;38(2):163–171.

55. El Assar M, Alvarez-Bustos A, Sosa P, Angulo J, Rodriguez-Manas L. Effect of Physical Activity/Exercise on Oxidative Stress and Inflammation in Muscle and Vascular Aging. Int J Mol Sci. 2022;23(15).

56. Nimmo MA, Leggate M, Viana JL, King JA. The effect of physical activity on mediators of inflammation. Diabetes Obes Metab. 2013;15 Suppl 3:51–60.

57. Barrea L, Di Somma C, Muscogiuri G, et al. Nutrition, inflammation and liver-spleen axis. Crit Rev Food Sci Nutr. 2018;58(18):3141–3158.

58. Di Giosia P, Stamerra CA, Giorgini P, Jamialahamdi T, Butler AE, Sahebkar A. The role of nutrition in inflammaging. Ageing Res Rev. 2022;77:101596.

59. Rocha VZ, Libby P. Obesity, inflammation, and atherosclerosis. Nat Rev Cardiol. 2009;6(6):399–409.

60. Saltiel AR, Olefsky JM. Inflammatory mechanisms linking obesity and metabolic disease. J Clin Invest. 2017;127(1):1–4.

61. Colpani V, Baena CP, Jaspers L, et al. Lifestyle factors, cardiovascular disease and all-cause mortality in middle-aged and elderly women: a systematic review and meta-analysis. Eur J Epidemiol. 2018;33(9):831–845.

62. Han H, Cao Y, Feng C, et al. Association of a Healthy Lifestyle With All-Cause and Cause-Specific Mortality Among Individuals With Type 2 Diabetes: A Prospective Study in UK Biobank. Diabetes Care. 2022;45(2):319–329.

63. Pai JK, Hankinson SE, Thadhani R, Rifai N, Pischon T, Rimm EB. Moderate alcohol consumption and lower levels of inflammatory markers in US men and women. Atherosclerosis. 2006;186(1):113–120.

64. Minzer S, Losno RA, Casas R. The Effect of Alcohol on Cardiovascular Risk Factors: Is There New Information? Nutrients. 2020;12(4).

65. Bielecka-Dabrowa A, Ebner N, Dos Santos MR, Ishida J, Hasenfuss G, von Haehling S. Cachexia, muscle wasting, and frailty in cardiovascular disease. Eur J Heart Fail. 2020;22(12):2314–2326.

66. Dev R, Bruera E, Dalal S. Insulin resistance and body composition in cancer patients. Ann Oncol. 2018;29(suppl_2):ii18–ii26.

67. Nakajima K, Yamaoka H, Morita K, et al. Elderly people with low body weight may have subtle low-grade inflammation. Obesity (Silver Spring). 2009;17(4):803–808.

